# SARS-CoV-2 Spike protein binding of glycated serum albumin - its potential role in the pathogenesis of the COVID-19 clinical syndromes and bias towards individuals with pre-diabetes/type 2 diabetes & metabolic diseases

**DOI:** 10.1101/2021.06.14.21258871

**Authors:** Jason K IIes, Raminta Zmuidinaite, Christoph Sadee, Anna Gardiner, Jonathan Lacey, Stephen Harding, Jernej Ule, Debra Roblett, Ray K Iles

## Abstract

Since the immune response to SARS-CoV2 infection requires antibody recognition of the Spike protein, we used MagMix, a semi-automated magnetic rack to reproducibly isolate patient plasma proteins bound to a pre-fusion stabilised Spike and nucleocapsid proteins conjugated to magnetic beads. Once eluted, MALDI-ToF mass spectrometry identified a range of immunoglobulins, but also in Spike protein magnetic beads we found a high affinity for human serum albumin. Careful mass comparison revealed a preferential capture of advanced glycation end product (AGE) glycated human serum albumin by the pre-fusion Spike protein.

The ability of bacteria and viruses to surround themselves with serum proteins is a recognised process of immune evasion. A lower serum albumin concentration is a reported feature of COVID-19 patients with severe symptoms and high probability of death. This binding preference of the Spike protein for AGE glycated serum albumin may contribute to immune evasion and influence the severity & pathology of SARS-COV2 towards acute respiratory distress. Thus, it can be hypothesised, contributing to the symptom severity bias and mortality risk for the elderly and those with (pre)diabetic and atherosclerotic/metabolic diseases who contract SARS-CoV2 infections.

**Graphic abstract:** 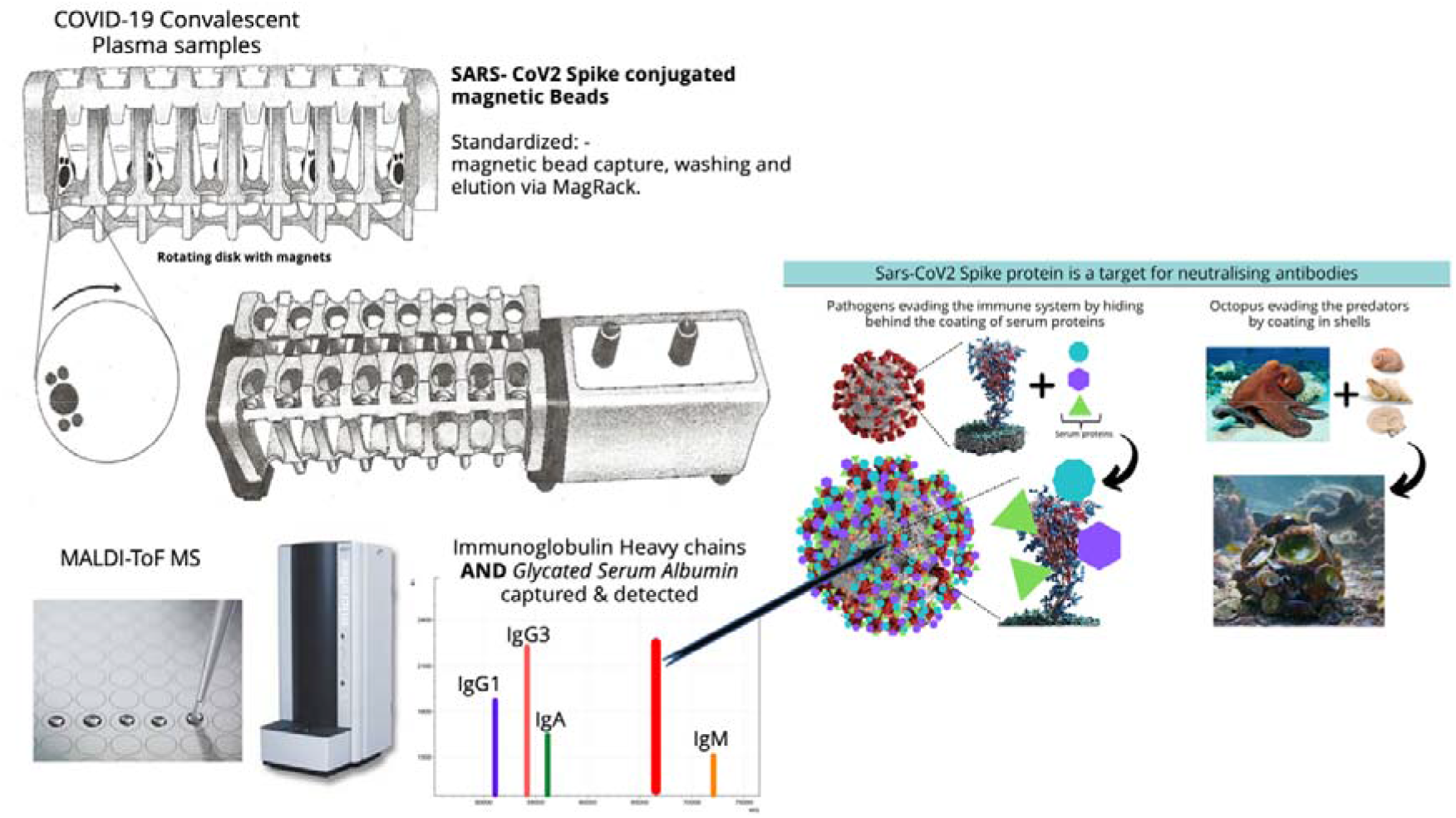

## Introduction

The disease syndrome COVID-19 is caused by the enveloped corona virus SARS-CoV2. Clinical presentation is varied from asymptomatic or mild to severe and life threatening. Antibodies to SARS-CoV2 envelope proteins are seen in those infected and are particularly elevated in those with severe illness [1].

The initial infection of the nasopharyngeal epithelia man be asymptomatic and the local immune response, such as mucosal IgA, in many, particularly the young, will be sufficient for the virus not to progress. However, if it does progress to involvement of the upper airways, the disease manifests with symptoms of fever, malaise and dry cough. There is a greater immune response during this phase involving the release of C-X-C motif chemokine ligand 10 (CXCL-10) and interferons (IFN-β and IFN-λ) from the virus-infected cells [2]. The majority of patients do not progress beyond this phase as the mounted immune response is sufficient to contain the spread of infection.

The virus-laden pneumocytes release many different cytokines and inflammatory markers such as interleukins, tumour necrosis factor-α, Interferons, monocyte chemoattractant protein-1 and macrophage inflammatory protein-1α. This ‘COVID-19 cytokine storm’ acts as a chemoattractant for neutrophils, CD4 helper T cells and CD8 cytotoxic T cells. Sequestered into the lung tissue, these cells are responsible for the subsequent inflammation and lung injury, culminating in an acute respiratory distress syndrome. [3,4].

At admission, the majority of patients had lymphopenia and platelet abnormalities, elevated neutrophils, aspartate aminotransferase (AST), lactate dehydrogenase (LDH) along with these inflammatory biomarkers [5]. Medical imaging (CT or X-ray) frequently shows patients have bilateral pneumonia and pleural effusion occurs in only 10% of the patients [6]. Disease severity is associated with higher level of neutrophils, AST, LDH, and C-reactive protein (CRP) lower level of platelets [7,8] and albumin [9,10].

Sars-CoV2 is an enveloped virus in which its RNA genome held by a polymer of Nucleocapsid is encased within a lipid membrane from which several virally encoded proteins are embedded or protrude. Key amongst these is the Spike protein complex. The Spike complex is the principal target for neutralising antibodies as this viral envelope surface exposed structure is functionally responsible for the virus target and entry into cells via the ACE 2 receptor expressed on the target cells. The spike complex is a trimer of the large S protein which, after binding to the ACE-2 receptor, undergoes activation via a two-step protease cleavage: the first one for S protein priming at the S1/S2 cleavage site and the second cleavage for activation at a position adjacent to a fusion peptide within the S_2_ subunit [11,12,13,14]. The initial cleavage stabilises the S2 subunit at the attachment site and the subsequent cleavage activates the S protein causing conformational changes leading to viral and host cell membrane fusion [15].

Numerous studies have shown high levels of anti-Nucleocapsid and anti-Spike protein antibodies present in the blood of patients developing severe illness as a result of SARS-CoV2. The nature a composition of these antibodies in relations to development of acute respiratory distress syndrome (ARDS) is the focus of much research. As part of an investigation of serum/plasma binding proteins to SARS CoV2 antigenic proteins albumin was found to bound strongly to stabilised complete spike protein but not nucleocapsid protein nor control protein G. Characterised by MALDI-ToF mass spectrometry the albumin bound was found to vary in molecular mass consistent with advanced glycation end products (AGE).

## Materials and methods

### Samples

Serum and plasma samples were obtained from Health care workers (HCWs) and patients referred to the Royal Papworth Hospital for critical care. COVID-19 patients hospitalised during the first wave and as well as NHS healthcare workers working at the Royal Papworth Hospital in Cambridge, UK served as the exposed HCW cohort (Study approved by Research Ethics Committee Wales, IRAS: 96194 12/WA/0148. Amendment 5). NHS HCW participants from the Royal Papworth Hospital were recruited through staff email over the course of 2 months (20^th^ April 2020-10^th^ June 2020) as part of a prospective study to establish seroprevalence and immune correlates of protective immunity to SARS-CoV-2. Patients were recruited in convalescence either pre-discharge or at the first post-discharge clinical review. All participants provided written, informed consent prior to enrolment in the study. Sera from NHS HCW and patients were collected between July and September 2020, approximately 3 months after they were enrolled in the study [16].

Clinical assessment and WHO criteria scoring of severity for both patients and NHS HCW was conducted following the ‘COVID-19 Clinical Management: living guidance (https://www.who.int/publications/i/item/WHO-2019-nCoV-clinical-2021-1).

For cross-sectional comparison, representative convalescent serum and plasma samples from seronegative HCWs, seropositive HCW and convalescent PCR-positive COVID-19 patients were obtained. The serological screening to classify convalescent HCW as positive or negative was done according to the results provided by a CE-validated Luminex assay detecting N-, RBD- and S-specific IgG, a lateral flow diagnostic test (IgG/IgM) and an Electro-chemiluminescence assay (ECLIA) detecting N- and S-specific IgG. Any sample that produced a positive result by any of these assays was classified as positive. The clinical signs of the individuals from which the sample was obtained ranged from 0 to 7 according to the WHO classification described above. Thus, the panel of convalescent plasma samples (3 months post-infection) were grouped in three categories: and A) Seronegative Staff (n=30 samples). B) Seropositive Staff (n=31 samples); C) Patients (n=38 samples).

### Antigen coupled magnetic beads

The viral Spike protein (S-protein) is present on virions as a pre-fusion trimers with the receptor binding domain of the S1 region stochastically open or closed, an intermediary where the S1 region is cleaved and discarded, and the S2 undergoes major confirmation changes to expose and then retract its fusion peptide domain [17]. Here the S-protein was modified to disable the S1/S2 cleavage site and maintain the pre-fusion stochastic confirmation [18].

Protein-G coupled magnetic beads were purchased from Cytivia Ltd (Amersham Place, Little Chalfont, Buckinghamshire, UK). Recombinant Nucleocapsid and recombinant stabilized complete spike protein magnetic beads were made by Bindingsite Ltd (Birmingham, UK).

### Isolation of bound proteins with MagMix, a semi-automated magnetic rack

Consistency in processing of magnetic bead-coupled antigen-captured binding-proteins from plasma sample was achieved using the semi-automated magnetic rack produced by the Crick Institute, London, UK (see figure 1, panel 1) further details at bitomix.com. This instrument has a series of magnets on rotating discs set either side of the tube containing samples and magnetic beads. Rotation of the magnets not only pulls the antigen magnetic beads to one side, as is done by the commercially available static magnets, but also has a series of other automated functions. These include:

**Figure 1.**
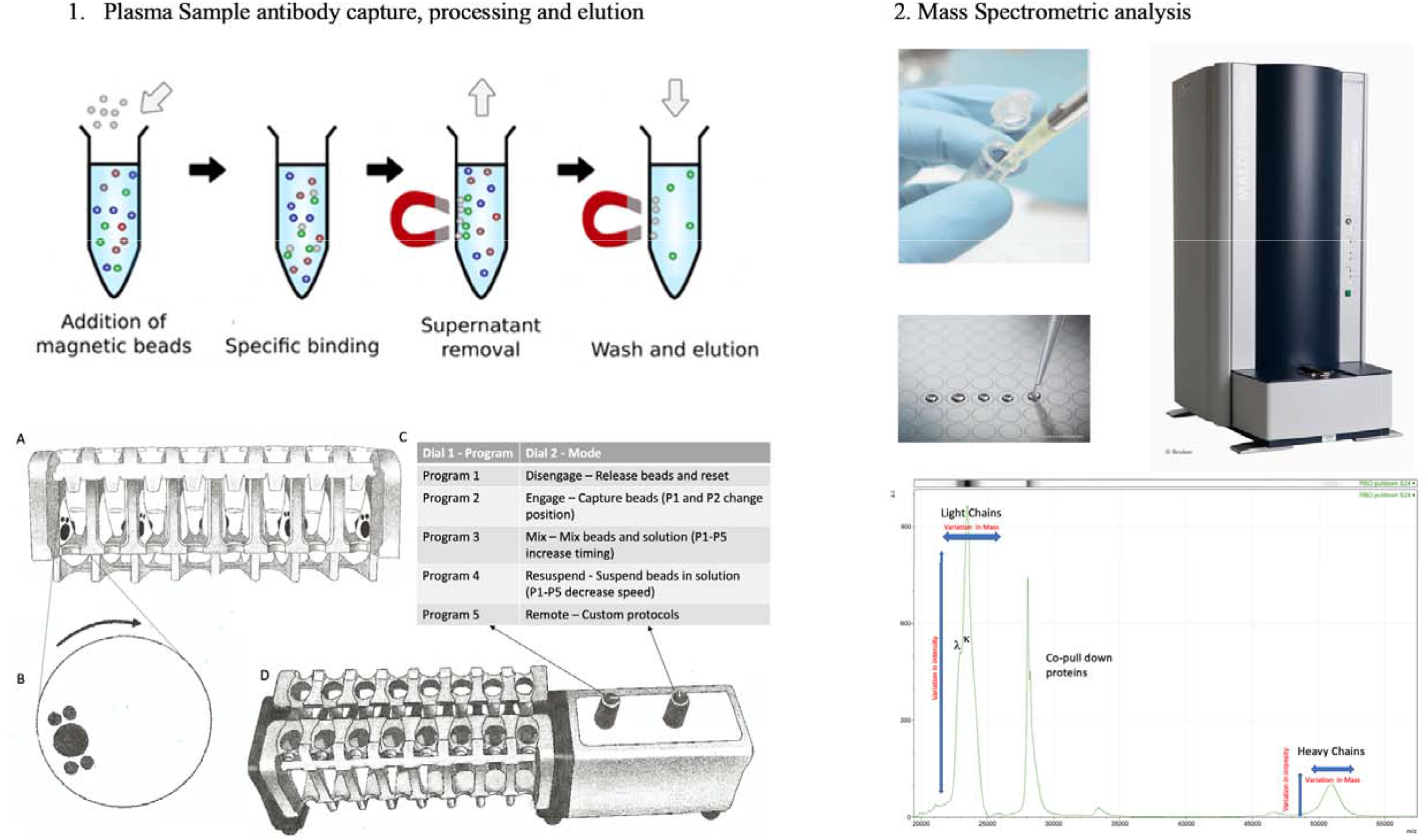
A schematic of the capture and elution of binding proteins such, as immunoglobulins, is illustrated in panel 1. Plasma samples are mixed with magnetic beads coupled with Nucleocapsid (N-Protein), stabilised complete Spike (Spike protein) or G-protein. The magnetic particle processing rack can simultaneously process sixteen samples in Eppendorf tubes (1A). Using rotating disk containing magnets (1B) either side of the sample allows capture, removal of denuded sample, mixing and efficient wash off of non-specific absorption and final elution of the capture binder proteins (1C and D) in a consistent standardised process. The eluted sample are spotted onto a MALDI-ToF mass spectrometry plate along with MALDI-matrix and subjected to mass spectral analysis following a standardised protocol and settings. Raw data is exported and analysed to give precise measurement of molecular mass of the eluted binder/complex proteins (e.g. Immunoglobulin light and heavy chains) along with intensity measurement for relative quantification purposes (Panel 2).

1. Engage/Disengage: Beads are immobilized allowing liquid to be aspirated or disengaged to transfer beads in solution to a new test tube.
2. Mix: Beads are pulled from one side of the tube to the other under a changing magnetic field in a gentle but thorough manner, enabling a repetitive and consistent washing.
3. Resuspend: Beads are suspended into liquid via a rapidly changing magnetic field and remain in liquid upon magnets disengaging after three cycles, beads may now be further processed or transferred to a new tube.
4. Remote: Beads can are washed and agitated via a custom protocol to be uploaded through the serial port.

To prepare the beads, empty 1.5µl microcentrifuge tubes were loaded into the automated magnetic rack. The rack was turned on and set to the ‘Disengage’ setting (Figure 1c). Protein G (GE), purified nucleocapsid or purified stabilized complete spike magnetic beads (Bindingsite, Birmingham UK) in their buffer solutions were vortexed to ensure an even distribution of beads within the solution, and 10µl of the appropriate magnetic beads were pipetted into each tube. 100µl of wash buffer, 0.1% Tween 20 in Dulbecco’s phosphate, buffered saline (DPBS) was then added into each tube before the rack was set to the ‘Resuspend’ setting. During this period, the magnets continuously circulated for several rotations before stopping. Once the rack had finished resuspending, the settings were changed to ‘Mix’ for several minutes, and then changed back to the ‘Resuspend’ setting to mix once more. The rack was set to ‘Engage’ allowing the wash buffer to be carefully discarded using a pipette, while taking care not to disturb the beads. The wash cycle was repeated three times. After 3 wash cycles, all of the remaining wash buffer in the tubes was discarded without disturbing the beads. The rack was set into the ‘Disengage’ setting.

To process the samples and elute the binding fraction from magnetic beads, 45µl of 10X DPBS was pipetted into each of the tubes containing the washed magnetic beads. 5µl of vortexed 1:10 diluted plasma was pipetted and pump mixed into a tube containing the beads, repeating for each plasma sample. The rack was set to the ‘Resuspend’ setting to ensure a thorough mix of the beads with the diluted plasma. After the resuspension, the rack was set to ‘Mix’ for 20 minutes. After the mix, the rack was set to ‘Engage’ and the remaining solution was carefully discarded as to not disturb the beads. A further 3 wash cycles were conducted using 0.1% DPBS. Subsequently, another 3 wash cycles were conducted after this, using ultra-pure water, discarding the water after the last cycle. 15µl of recovery solution (20mM tris(2-carboxyethyl)phosphine (TCEP) (Sigma-Aldrich, UK) + 5% acetic acid + ultra-pure water) was pipetted into the tubes. The tubes were removed from the rack and mixed by flicking the tube to fully disturb the beads. All of the liquid was sent to the bottom of the tube by firmly tapping the tube on a surface to bring the droplets down. The tubes were securely placed back into the rack and were run alternatively between the ‘Resuspend’ setting and the ‘Mix’ setting for 5 minutes. The rack was set to ‘Engage’ and the recovery solution was carefully removed using a pipette and placed into a clean, labelled 0.6µl microcentrifuge tube. This recovery solution was the eluant from the beads and contained the desired proteins.

### Sample Analysis by MALDI-ToF Mass spectrometry

Mass spectra were generated using a 15mg/ml concentration of sinapinic acid (SA) matrix. The elute from the beads was used to plate with no further processing. 1µl of the eluted samples were taken and plated on a 96 well stainless-steel target plate using a sandwich technique. The MALDI-ToF mass spectrometer (microflex® LT/SH, Bruker, Coventry, UK) was calibrated using a 2-point calibration of 2mg/ml bovine serum albumin (33,200 m/z and 66,400 m/z) (Pierce™, ThermoFisher Scientific). Mass spectral data were generated in a positive linear mode. The laser power was set at 65% and the spectra was generated at a mass range between 10,000 to 200,000 m/z; pulsed extraction set to 1400ns.

A square raster pattern consisting of 15 shots and 500 positions per sample was used to give 7500 total profiles per sample. An average of these profiles was generated for each sample, giving a reliable and accurate representation of the sample across the well. The raw, averaged spectral data was then exported in a text file format to undergo further mathematical analysis.

### Spectral Data processing

Mass spectral data generated by the MALDI-ToF instrument were uploaded to an open-source mass spectrometry analysis software mMass™ [19], where it was processed by using; a single cycle, Gaussian smoothing method with a window size of 300 m/z, and baseline correction with applicable precision and relative offset depending on the baseline of each individual spectra. In software, an automated peak-picking was applied to produce peak list which was then tabulated and used in subsequent statistical analysis.

### Statistical analysis

Peak mass and peak intensities were tabulated in excel and plotted in graphic comparisons of distributions for each antigen capture and patient sample group. Means and Medians were calculated and, given the asymmetric distributions found, non-parametric statistics were applied, such as Man Whitney U test, when comparing differences in group distributions.

## Results

This study relies on use of antigen-coupled magnetic beads to capture human serum proteins, which requires robust and reproducible processes of magnetic bead capture, washing, agitation and target binding protein elution (Figure 1c-d). If this is performed manually, the result can vary dramatically due to variations in timings between steps, in the intensity of mixing, while insufficient washing can result in large amounts of non-specific binding proteins being recovered and lead to variable efficiency of target binding proteins recoveries. To overcome these problems, and to minimise individual operator variability, we employed a newly-developed automated magnetic rack system. By enabling automation, this system leads to consistent timing and intensity of binding, mixing and elution steps, and ensures gentle but thorough washing by enabling longer mixing with more repetitions.

Post elution from respective antigen coupled magnetic beads MALDI-ToF mass spectra was obtained and peaks recorded. These were matched against reference MALDI-ToF mass spectra of preparation of purified human serum proteins run under the same reducing and acetic acid pH conditions: human serum albumin, Transferrin (Merck Life Science UK Ltd, Dorset, England), IgG1, IgG3, IgA & IgM (Abcam, Discovery Drive, Cambridge, Biomedical Campus, Cambridge, UK).

Although immunoglobulin light chains and heavy chains were recovered as expected (and subject to detailed analysis reports elsewhere) there was a significant recovery of human serum albumin by the magnetic beads to which stabilised complete Spike was conjugated (Figure 2). Indeed the recovery was conspicuous in this one magnetic bead coupled SAR-S-Cov2 protein series recovered levels of HSA to a level similar to that seen for Protein G binding and recovery of human IgG1, to which it has a known specificity. The same samples failed to bind albumin to the same degree despite its vast abundance in the samples to Protein-G or Nucleocapsid (see figure 2). Human serum albumin was detected in 72% of the samples of the Spike protein conjugated magnetic beads (compared to 33% for nucleocapsid and 13% Protein-G), and at an intensity level over 10x higher than that found for residual non-specific binding of HSA for nucleocapsid and Protein-G (mean values; 1923 AU Spike protein, 159 AU N-protein and 100 AU Protein-G).

**Figure 2.**
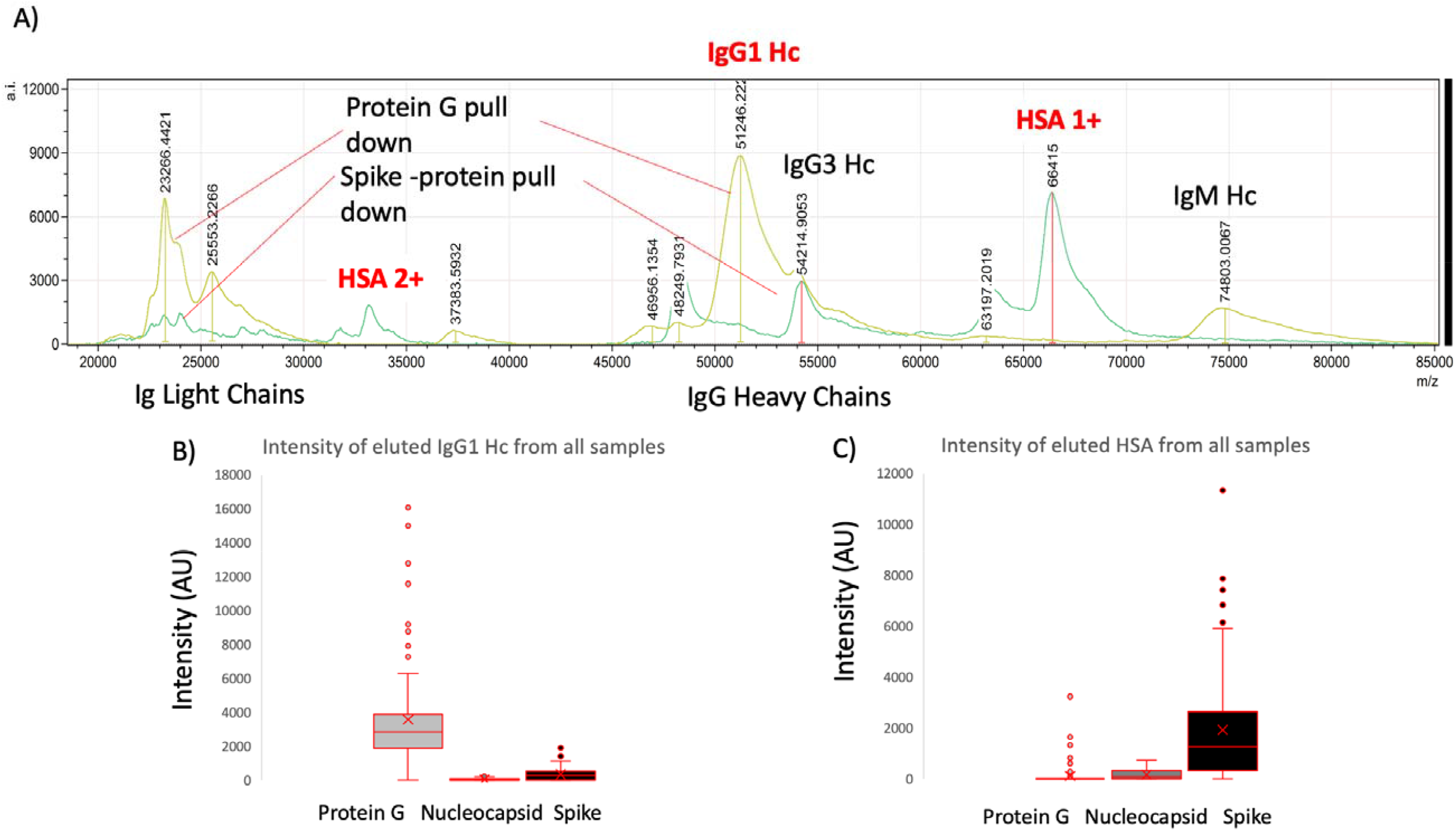
Example mass spectra of eluted sample from protein G bead captured proteins overlaid with Spike protein captured plasma proteins (panel A) and relative intensities of IgG1 heavy chains (IgG 1 Hc) recovered from the same samples by Protein G, nucleocapsid and stabilised spike protein (panel B); versus HSA (single: 1+ and doubly charged: 2+) recovered from the same samples by Protein G, nucleocapsid and stabilised spike protein (panel C).

Closer examination of the bound and eluted HSA masses revealed that the average mass of the albumin recovered by the stabilised complete Spike protein was higher than that found for the low level non-specific absorption to nucleocapsid and Protein G (Δ152m/z, p<0.05, see figure 3 panel A). The binding of this higher molecular mass albumin was not a specific unique feature of plasma from patients who had recovered from COVID19 ARDS; but was also being bound from sero-negative and sero-positive HCW samples (see figure 3 panel B). The average increase in HSA mass was 152m/z but ranged from ranged from 50 up to 500+m/z and Spike protein affinity was greatest for albumin with increased mass of about 150m/z (see figure 3 panel B).

**Figure 3.**
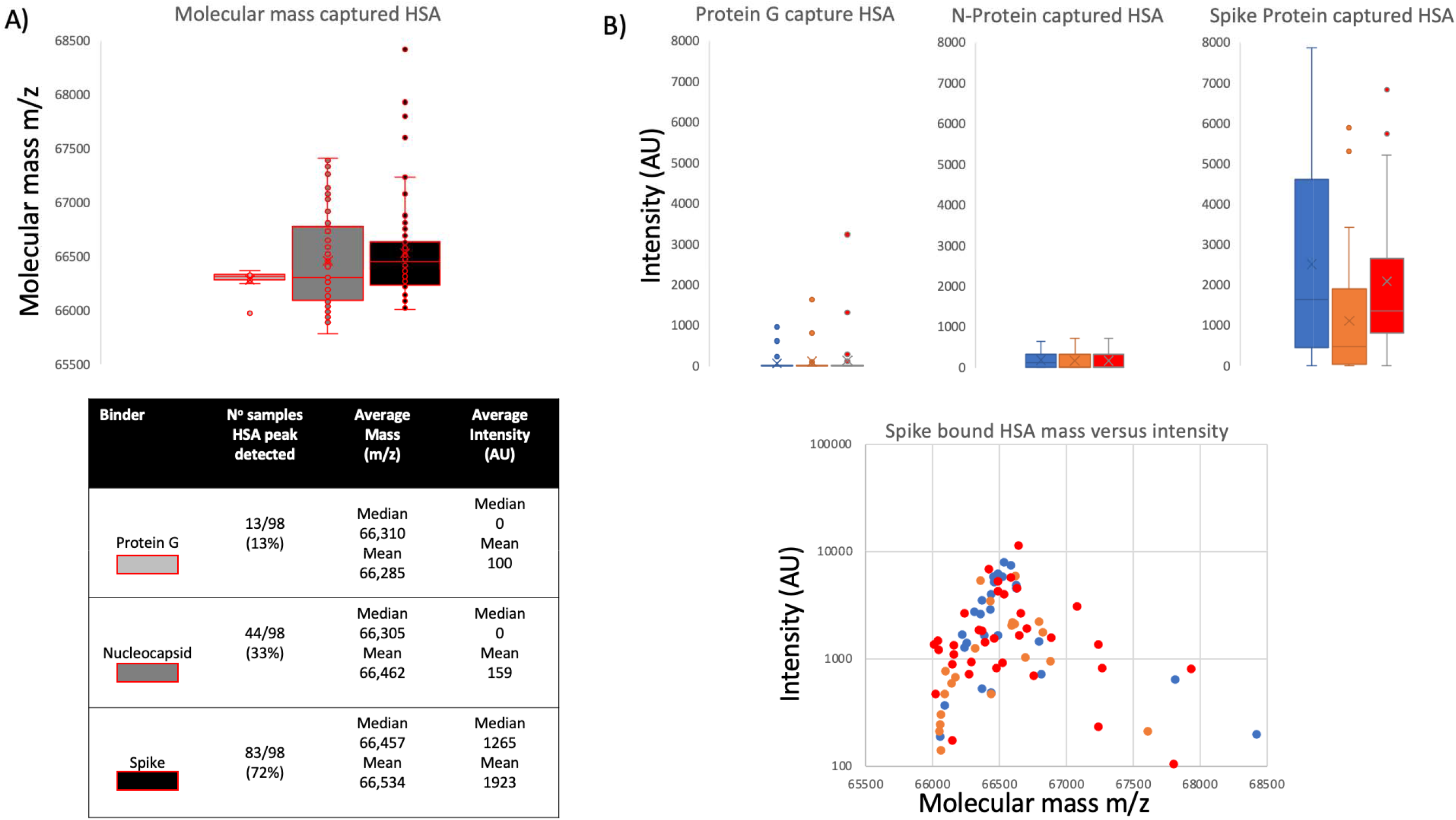
Distribution of mass determined for captured and eluted human serum albumin (HSA) and the relative intensity of the increased mass albumins being bound: Panel A illustrates the mass variance of Protein G, Nucleocapsid (N-Protein) and stabilised complete Spike Protein along with average intensity. Panel B illustrates a lack of differences between patient groups plasma samples HSA binding by protein G, Nucleocapsid and stabilised complete Spike protein; whilst a clear preference of the S-protein to bind a higher molecular weight albumin from all samples. Blue represents data from SARS-CoV2 sero negative HCW, Orange from SARS-CoV2 HCW sero-positive and having recovered from with mild symptoms and Red sample data from convalescent patients recovering from COVID-19 ARDS.

## Discussion

By far the greatest at-risk group from dying following infection with SARS-CoV2 are those over 60 who developed COVID-19 symptoms; accounting for nearly 95% of all such deaths. Male sex, and comorbidities such as y are hypertension, obesity and type II diabetes are also risk factors for severe disease[20,21,22,23].

The underlying pathological mechanism of these associations is unknown, but is likely to be multifactorial rather than a single determinant/marker [24].

Strong and specific human serum albumin binding to magnetic beads to which stabilised prefusion Spike protein is conjugated was found to occur with all plasma samples; be it from sero-negative & sero-positive medical staff (who had no symptoms or mild symptoms) or COVID-19 patients who had been treated on for ARDS on COVID-19 ITU wards. Albumin in these same plasma samples was not found to bind strongly to nucleocapsid or Protein-G coated magnetic beads, which acted as a control. This is despite the fact that significant human serum albumin binding has been reported as a feature of Protein-G [25].

Many microorganisms that cause seriously harmful infections have developed mechanisms to evade the immune system. One such mechanism includes the specific binding of serum proteins to mask antigenic sites and thus evade antibody neutralisation and other immune processes. The spike protein is an extremely large antigenic target projecting and exposed from the virion Envelope. It is the focus of neutralising antibodies [26]. Thus, the ability to evade neutralising antibodies by coating with abundant serum protein(s), until such time as target cell receptor binding has occurred, is an ideal strategy to evolve, particularly when entering the blood stream. Bacteria express families of genes coding for surface proteins that bind serum proteins, in particular those that bind immunoglobulins, i.e. IgG by Protein-G (human group C and G streptococci) and Protein-A (*Staphylococcus aureus)*. An analogous gene to that which codes for Protein-G has been found to code for a protein, termed PAB, which binds human serum albumin. PAB is highly expressed by the anaerobic bacterium *Peptostreptococcus magnus* [25] and such expression has been correlated with virulence of *P*.*magnus* strains.

Similarly, the viral nano-particle surface can absorb host proteins as a coat or protective corona and this in turn affect the hosts physiology and immune response [27,28,29]. For example respiratory syncytial virus (RSV) and herpes simplex virus type 1 (HSV-1) accumulate a rich protein corona from the biological fluids, and this protein corona affected viral infectivity and immune cell activation [29]. Recent molecular modelling has indicated that the S1 subunit of the Spike protein would bind HSA in such a way that it blocked access to antigenic sites on the RBD [30]. Thus even weak binding affinity for HSA could provide a virulence enhancement via some protection/evasion from RBD neutralisation antibodies.

Since detection of the eluted bound albumin was via MALDI-ToF mass spectrometry a more precise molecular mass of the bound albumin could be determined. It was found that the molecular mass of the elevated levels of albumin binding to the stabilised spike protein was higher than expected; and higher in molecular mass than that of the low levels of non-specific bound albumin which could be detected in some elution samples from nucleocapsid and Protein-G coated magnetic beads (Figure 3).

Viruses such as influenza bind to specific glycan as part of their attachment and invasion mechanisms e.g. the viral envelope protein haemagglutinin recognises and binds sialic acid glycan residues [31]. Albumin is not a glycosylated protein [32]; however, the increases in molecular mass seen here correspond to known advanced glycation end products (AGE) which form from blood saccharide reaction with long lived and abundant serum proteins. The AGE reaction occurs on specific lysine and arginine amino acid side chains of human serum albumin (see figure 4). Glycation of serum proteins is more rapid and abundant than glycation of haemoglobin and has been used as biomarker of both diabetes and arthrosclerosis [33].

**Figure 4.**
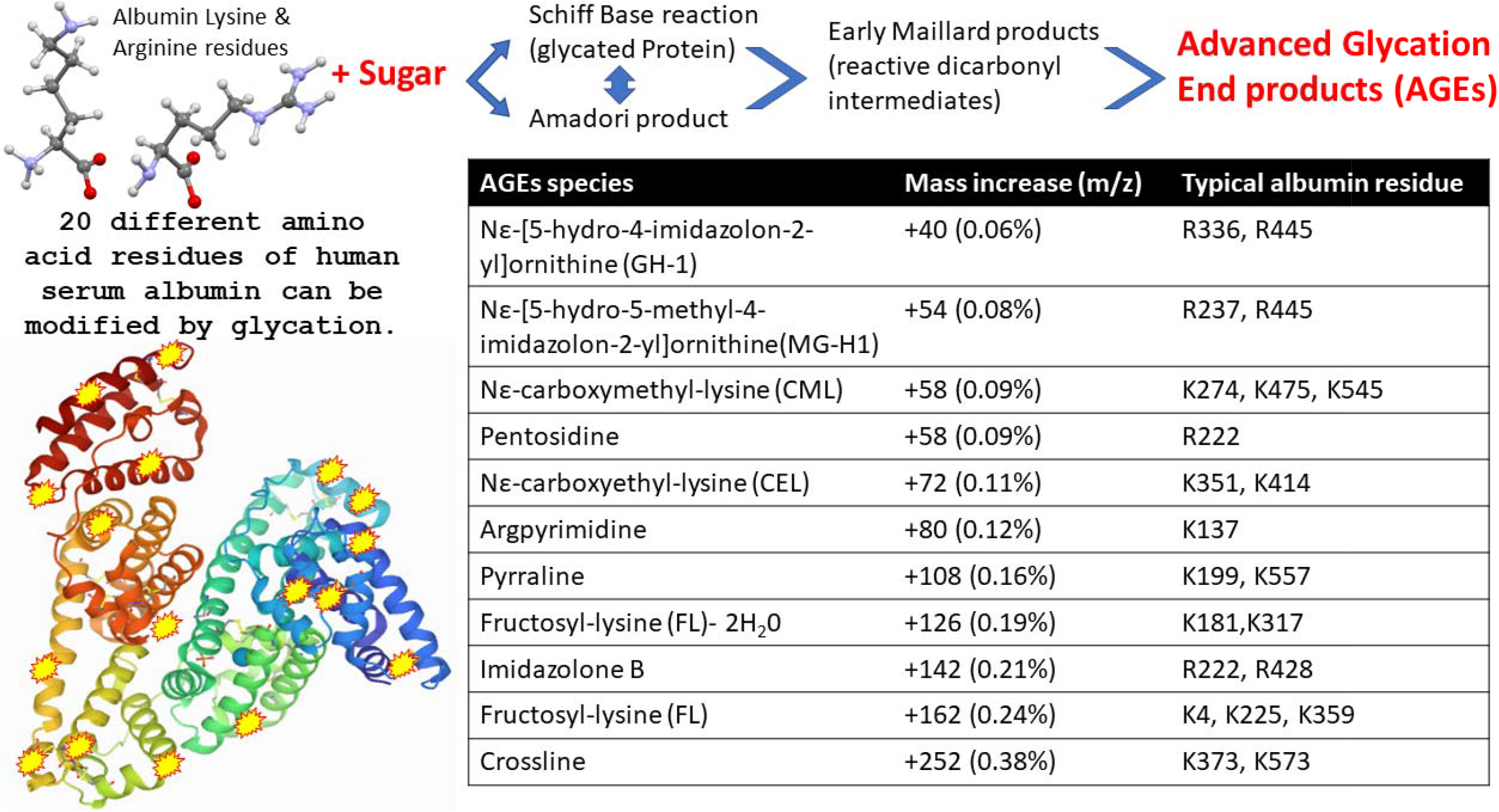
Illustration of advanced glycation end product (AGE) as a result of human serum albumin Schiff base/ Amadori/Maillard reactions with blood circulating reducing monosaccharides (glucose and fructose), resulting in modification to exposed Lysine and Arginine amino acid side chains. The molecular mass effects of these AGE reactions on specific HSA residues is also detailed.

Paradela-Dobarra *et al* [34] demonstrated that Advanced Glycation End product (AGE) modification of Albumin could be detected by direct MALDI-ToF mass spectrometry via an increase in the averaged albumin peak molecular mass. This was reflected in a ratio consistent increase in mass of the albumin spectral singly and doubly charged peaks. We also detected similarly increased average mass of, MALDI-ToF MS determined singly and doubly charged, HSA peaks in our samples and the results are consistent with AGE glycation forms of HSA being preferentially captured by Spike protein.

A stronger affinity to glycated/AGE’d serum albumin would enhance such a virulence effect of corona virus in those with pre-diabetes and metabolic diseases.

### Potential clinical correlations

An unusual feature of the COVID-19 infection disease is micro thrombosis and localised disruption of the osmotic potential with pulmonary micro vascular dilation. Pulmonary thrombosis is common in sepsis□induced ARDS. Coagulation dysfunction appears to be common in COVID□19, and is detected by elevated D□dimer levels. In fatal cases there is diffuse microvascular thrombosis, suggesting a thrombotic microangiopathy, and most deaths from COVID□19 ARDS have evidence of thrombotic disseminated intravascular coagulation [35](Wang et al., 2020). This may explain some of the atypical or unexpected manifestations seen in the lung, such as dilated pulmonary vessels on chest CT, and episodes of pleuritic pain. Vascular enlargement is rarely reported in typical ARDS, yet was seen in most cases of COVID□19 ARDS.

Low circulating serum albumin is associated with pleural effusion, oedema and vascular constriction, as a result of serum albumin being leaking or being sequestered within the interstitial tissues. However, in COVID-19 ARDS we see a lower overall serum albumin and yet pulmonary vascular dilation. Thus, where is the serum albumin being lost to?

The binding of albumin by SARS-CoV-2 has been postulated by Johnson *et al* [37] as a molecular contributor to fluid tissue-vascular imbalance giving rise to septic shock in COVID-19 cases. The possibility is that it is being caught locally within the pulmonary vasculature and contributing to the pathologic mechanism of local vascular dilation e.g. within micro-thrombolytic clots found in the pulmonary blood vessels [38,39]. Furthermore, deposition of glycated/AGE modified albumin has also been correlated with hyperinflammatory process seen in cardiovascular disease [34].

As demonstrated by use of antigen-coupled magnetic beads via robust and reproducible processes of magnetic bead capture, washing, agitation and target binding protein elution; the stabilised/prefusion spike protein has a binding affinity for human serum albumin, in particular glycated/AGE’d albumin. The correlation of type 2 diabetes, obesity and age with greater probability of suffering from ARDS as a result of SARS-CoV2 infection that persist and progress; may involve molecular features of the Spike complex that include its higher affinity for glycated/AGE’d serum albumin. A potential hypothesis is that the ability to evade immune responses is by hiding behind a coating of serum proteins, much like an octopus grabs shells and stones to hide from predatory sharks (Blue planet II 2017) [40]. However, in so doing it may cause tissue-vascular fluid physiological changes seen in COVID-19 ARDS as a result of the proteins it has bound and then deposits, particularly in those with subclinical pre-diabetes and metabolic diseases.

## Data Availability

Data will be made available upon request

## Abbreviations

HCW: Health care workers
ARDS: Acute respiratory distress syndrome
ITU: Intensive Therapy unit
AGE: advanced glycation end products
MALDI-ToF: Matrix assisted laser desorption ionization – time of flight.
HSA: Human serum albumin

## Acknowledgements

This study was undertaken by the Humoral Immune Correlates to COVID-19 (HICC) consortium, funded by the UKRI and NIHR; grant number G107217 (COV0170 - HICC: Humoral Immune Correlates for COVID-19). RKI is also funded by NISAD Ideell Förening (charitable association) Organisationsnummer 802528-6157

We grateful acknowledge the loan of a prototype magnetic bead processing rack developed at the Francis Crick Institute London, UK, Bruker UK Ltd, Coventry for the loan of Microflex MALDI-Tof mass spectrometer and Dr Erika Tranfield and Dr Julie Green for technical support with running and data export from the Bruker Microflex.

## Notes

### Competing Interest Statement

The authors have declared no competing interest.

### Clinical Trial

Not a clinical trial

### Author Declarations

Study approved by Research Ethics Committee Wales, IRAS: 96194 12/WA/0148. Amendment 5

### Summary of Updates

Changes of text to reflect limitations on clinical emphasis and put emphasis more on biochemistry findings and methodology (wrong revision was uploaded by mistake)

